# REM sleep infraslow rhythms correlate with neuropsychiatric symptom severity

**DOI:** 10.64898/2026.10.06.26364874

**Authors:** Lezio S. Bueno-Junior, Peyton T. Wickham, Carolyn E. Jones-Tinsley, Miranda M. Lim, Brendon O. Watson

## Abstract

**Study objectives:** Rapid eye movement (REM) sleep is commonly divided into phasic and tonic substates based on higher and lower oculomotor activity, respectively. Our previous study described an alternative framework in which REM temporal structure forms a continuum of infraslow rhythms (∼0.01 Hz) across brain and body physiology. We now ask whether these infraslow rhythms vary with neuropsychiatric symptoms, given the long-standing but still inconsistent links between REM and mood- or trauma-related disorders.

**Methods:** Using a cross-sectional polysomnographic dataset from male veterans, we examined 59 participants for correlations between REM infraslow rhythms and neuropsychiatric symptom scores, excluding participants with moderate/severe apnea.

**Results:** Oscillations in REM respiratory rate, but not cardiac rate or electroencephalogram beta power, showed higher infraslow frequencies in participants with multiple symptoms, including posttraumatic stress, pain, anxiety, depression, and cognitive difficulties. No significant correlations were found between these symptoms and conventional REM metrics (e.g., REM density, latency and duration). To further examine REM beyond phasic/tonic binarization, we analyzed the timing between oculomotor activity and infraslow oscillations. Eye movements occurred preferentially during periods of high respiratory rate, low beta power and intermediate cardiac rate, with no changes in these temporal relationships across symptom severity.

**Conclusions:** Neuropsychiatric associations with REM infraslow rhythms spanned multiple symptom domains but were physiologically specific, with respiratory rate oscillations representing the strongest readout of symptom burden. Furthermore, REM infraslow cycles across brain and body were differentially aligned with eye movements: a temporal pattern that may serve as a physiological target for neuropsychiatric and mechanistic research.

**Statement of significance:** Rapid eye movement (REM) sleep has long been studied in relation to neuropsychiatric conditions, but traditional measures of the timing and duration of REM episodes have shown limited consistency for symptom tracking. This study addresses this gap by highlighting a less explored physiological feature within REM episodes: minute-scale, or infraslow, rhythms in respiratory and cardiac rate, as well as oculomotor and brain activity. We report that REM infraslow oscillations, particularly in respiratory rate, occur at higher frequency in participants with worse trauma/mood symptoms and cognitive difficulties. This work is evidence of a possible new REM sleep-based biomarker for multiple diseases.

## Introduction

Sleep is bidirectionally linked to brain health, both influencing and reflecting neurological and behavioral dysfunctions. To better understand these associations, researchers and clinicians have conventionally discretized polysomnographic (PSG) recordings into fixed-duration periods (e.g., 30-s epochs) labeled as WAKE, non-rapid eye movement sleep subtypes (NREM1-3), or rapid eye movement sleep (REM) [1]. However, a growing body of research has investigated physiological variations within these states and at finer temporal resolutions [2–11], revealing a deeper level of sleep structure with potential relevance to disease monitoring [1].

REM temporal structure is an important frontier in this regard, as research into its relationship with multiple diseases is still at an early stage. REM is commonly divided into two substates, phasic and tonic, respectively defined by higher and lower incidence of motor transients (e.g., rapid eye movements) [3], but also by differences in sleep fragility and brain activity [12–27]. However, the relevance of these REM substates, or their alternation, for health and disease remains poorly known. This reflects the lack of established metrics for quantifying the temporal organization of phasic and tonic REM. For example, REM density, a relatively well-known intrastate parameter [28], summarizes the incidence of eye movements, but does not capture when they occur or whether phasic/tonic features alternate with quantifiable periodicity.

Our previous work on REM sleep [29] described an approach to address this gap. Using both mouse electrophysiology and human PSG, we characterized intra-REM physiological variations recurring semi-regularly over tens of seconds to minutes (∼0.01 Hz): a frequency range known as infraslow [30]. In human PSG, we quantified REM infraslow oscillations in respiratory rate, cardiac rate, and electroencephalogram (EEG) beta power using power spectral density analysis [29]. Furthermore, using rapid eye movements as a temporal reference, we found that these REM infraslow oscillations show distinct temporal alignments to oculomotor activity, with eye movements occurring preferentially in periods of high respiratory rate, low beta power, and intermediate cardiac rate [29].

Thus, our previous work suggests that the binary phasic/tonic REM framework is incomplete without considering underlying physiological oscillations, which are quantifiable in terms of infraslow periodicity and different relative timings across brain and body signals [29]. This comprehensive approach to REM could be relevant to sleep medicine and psychiatry, as REM has long been linked to symptoms related to emotional regulation, memory and trauma [31–33], yet conventional REM metrics (e.g., REM density, latency and duration) have shown inconsistent associations with these symptoms [28,34–40].

Building on this background, we asked whether REM infraslow rhythms carry information about sleep quality and mood- or trauma-related states rather than representing a clinically neutral feature of REM. While our previous work used PSG from nine healthy participants (in addition to experiments in mice) [29], the present study examined a separate PSG cohort of 59 human participants with self-reported measures of sleep quality and neuropsychiatric symptoms. We found that one particular metric, the infraslow frequency of REM respiratory rate oscillations, correlated with a group of symptoms, including posttraumatic stress disorder (PTSD), anxiety, depression, neurobehavioral symptoms, cognitive difficulties, insomnia, and pain intensity ratings, suggesting that specific infraslow rhythms within REM may track multimorbidity burden. These results are discussed in relation to REM temporal structure and clinical implications.

## Methods

### Overview of participant data: deidentification and curation

This study used retrospective analysis of a cross-sectional symptom survey combined with PSG in the same participants. Specifically, participants were referred for PSG at an American Academy of Sleep Medicine (AASM)-accredited sleep clinic, located at the Veterans Affairs Portland Health Care System. There, participants completed neuropsychiatric questionnaires followed by overnight PSG (**Figure 1A**). Symptom surveys and PSG recordings were then saved in a restricted-access database, with personal identifiers replaced by a 15-character file name (e.g., XE52XBRY4AS22IB; **Figure 1A**). These random alphanumeric codes served both to deidentify each participant and to prevent investigators from memorizing participant-symptom associations, thus protecting the study from bias.

**Figure 1.**
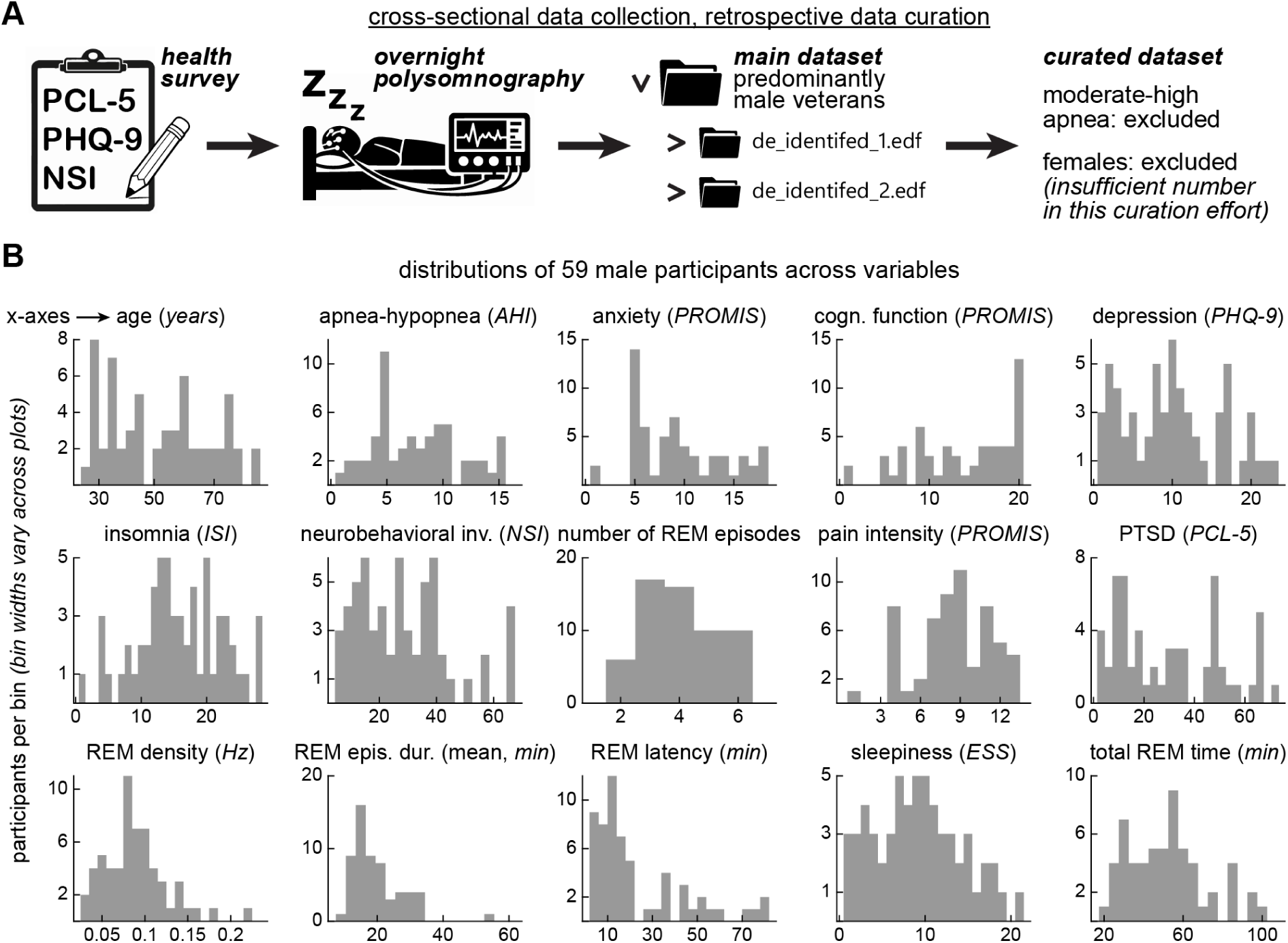
Overview of participant data: deidentification and curation. (A) Participants referred to an AASM-accredited sleep clinic (Veterans Affairs Portland Health Care System) answered neuropsychiatric and sleep quality surveys prior to overnight PSG, forming a deidentified dataset with >5,000 participants collected since 2015. The present retrospective analyses included a curated subset of 59 male participants with normal-to-mild apnea-hypopnea index scores. (B) Distributions of participants across analyzed variables, shown on their original scales. See **Table 1** for descriptive statistics and distribution normality tests. These raw distributions provide context for later analyses in which age, neuropsychiatric and sleep quality variables are transformed into derived metrics, including participant rankings and z-scores (Figures 3–5).

**Table 1.**
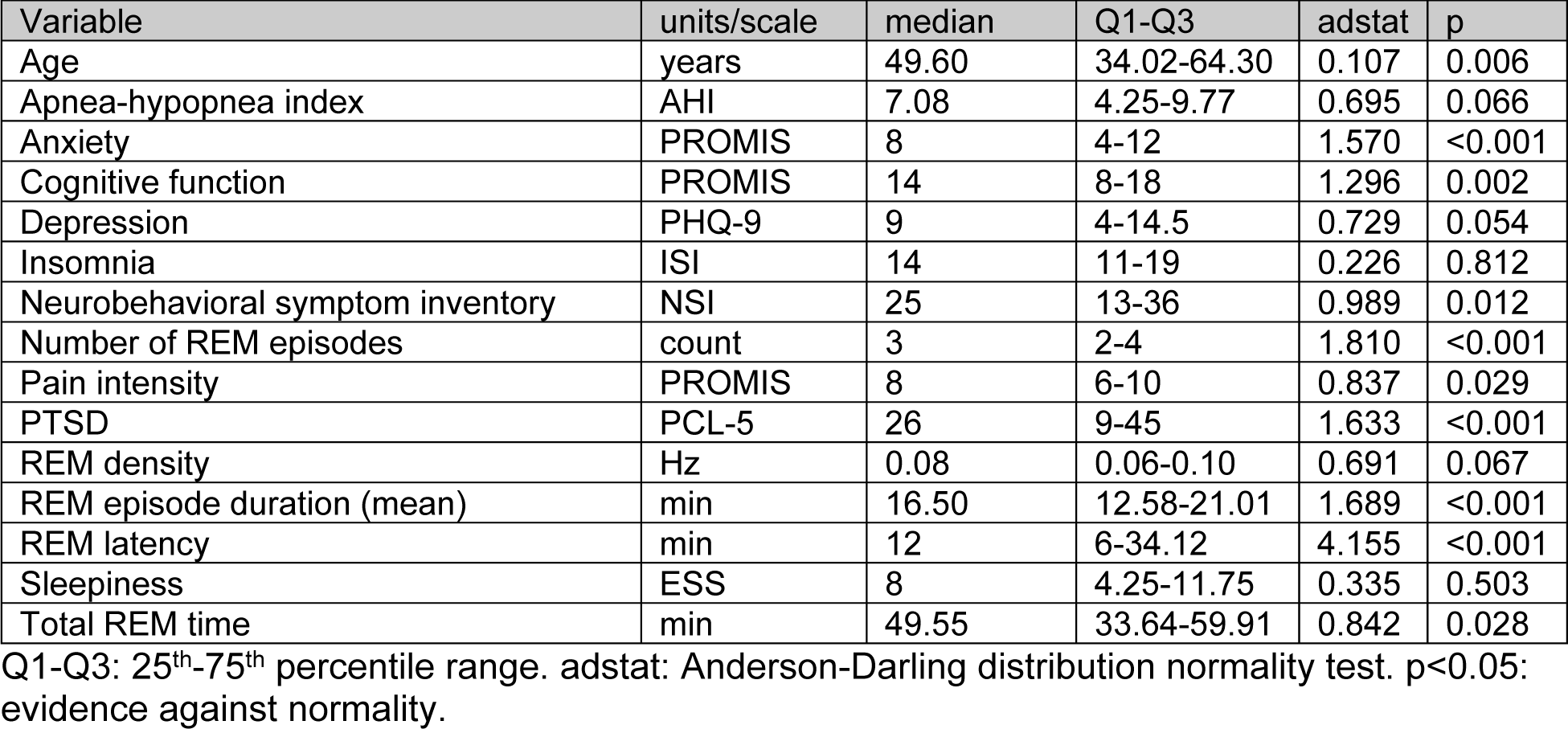
Summary statistics and normality tests for participant characteristics, symptom scores, and conventional REM sleep metrics (n = 59 male veterans).

Using these methods, a dataset with >5,000 participants has been constructed since 2015. For the current exploratory study, a subset of deidentified PSG files and associated symptom burden data from 59 participants was curated from the Sleep Laboratory at the VA Portland Health Care System, and then shared with the University of Michigan Medical School under an approved Data Use Agreement for the main analysis (see protocol numbers below). During this curation, only participants with an apnea-hypopnea index (AHI) lower than 15 (i.e., normal-mild range) were included. This AHI range was then retained as a variable for analysis (**Figure 1A-B**). Furthermore, only males were analyzed, reflecting their predominance in this veteran dataset (88% male). We acknowledge that sex differences in REM infraslow rhythms may exist, which is a direction for study in future work. Furthermore, no statistical power calculations were performed a priori as this was an exploratory, retrospective study. The sample size used here (n=59) reflects this curation effort, which was designed to provide a first-pass analysis of REM infraslow rhythms for neuropsychiatric research.

Regarding sleep quality and body movement criteria, participants were included if their PSG showed: (1) more than 6 h of sleep; (2) more than 20 min of REM sleep across two or more REM episodes; (3) periodic limb movement index of zero; (4) less than 5% of 30-s epochs with movement artifacts. This curation resulted in a diverse age range (24-84 years), which was also included among analyzed variables (**Figure 1B**), like the AHI range mentioned above. Distributions of age, AHI, and several other sleep quality and neuropsychiatric variables are illustrated in **Figure 1B** and statistically summarized in **Table 1**. These variables were examined for distribution normality, for correlations with one another, and for correlations with REM infraslow physiology metrics, as described later in this Methods section.

This noninterventional analytical study was conducted under Institutional Review Board protocols (VA Portland IRB #4108; University of Michigan IRB #HUM00281242) and a Data Use Agreement (#26-UFA00968) between the collaborating institutions, in accordance with ethical principles of the Belmont Report and the Declaration of Helsinki.

### Polysomnography

PSG consisted of a Type I, attended overnight recording using Polysmith equipment and software (Nihon Kohden, Japan) at 200-Hz sampling rate. EEG procedures followed the 10-20 electrode placement system, consisting of six scalp electrodes placed at F3, F4, C3, C4, O1, and O2. Other body physiology channels included bilateral electrooculography (EOG), electrocardiography (ECG), chin electromyography (EMG), abdominal/thoracic respiratory movement (plethysmography belts), airflow (nasal pressure transducer), and snoring (microphone). After each recording, an AASM-accredited technician manually scored sleep stages in 30-s epochs, according to the AASM manual [41]. Each 30-s epoch was scored as either WAKE, NREM 1-3, or REM. PSG and sleep scoring data collected at VA Portland were subsequently analyzed at the University of Michigan.

### Symptom surveys and basic REM metrics

In addition to age and AHI, eight symptom surveys were answered by each participant prior to PSG: anxiety (Patient-Reported Outcomes Measurement Information System, PROMIS [42]), cognitive function (PROMIS), pain intensity (PROMIS), PTSD (PTSD Checklist for DSM-5, or PCL-5 [43]), depression (Patient Health Questionnaire 9, PHQ-9 [44,45]), neurobehavioral symptom inventory (NSI [46,47]), insomnia severity index (ISI [48,49]), and sleepiness (Epworth Sleepiness Scale, ESS [50,51]) (**Figure 1B**). Moreover, five basic REM metrics were analyzed using MATLAB (MathWorks, Natick, MA, USA): number of REM episodes, total REM time (min), latency of first REM episode (min), mean REM episode duration (min), and REM density (here measured in Hz: number of eye movements divided by REM time) (**Figure 1B**). See below for details on eye movement detection. A link to the code repository is provided in the Data Availability section.

### REM epochs

Data processing in this and subsequent sections was also performed in MATLAB (see link to the code repository in the Data Availability section). For the illustration in **Figure 2A**, a representative overnight sleep recording was analyzed in its entirety, with emphasis on selected channels: an EEG montage (F3A2), left/right EOG, abdomen plethysmography, and ECG. The F3A2 montage was processed into a spectrogram (*spectrogram* function, 2-s window, 50% overlap, 2-51 Hz frequency range, 1-Hz frequency resolution, 120-s moving mean), z-scored across frequency bins and then z-scored across time bins. A beta-band power curve was then produced by averaging across 15-30 Hz frequency bins (**Figure 2A**). EOG was processed as the absolute difference between the left and right eye channels, thus magnifying eye movement events for timestamping using the *findpeaks* function (minimum peak prominence between 100-500, adjusted heuristically per participant) (**Figure 2A**). Respiratory cycles (inspiratory peaks in abdominal plethysmography) and heartbeats (R peaks in ECG) were also timestamped using the *findpeaks* function (same peak prominence method as above). These timestamps were then counted in 1-s bins using the *histcounts* function, producing respiratory and cardiac rate time series in Hz (**Figure 2A**).

**Figure 2.**
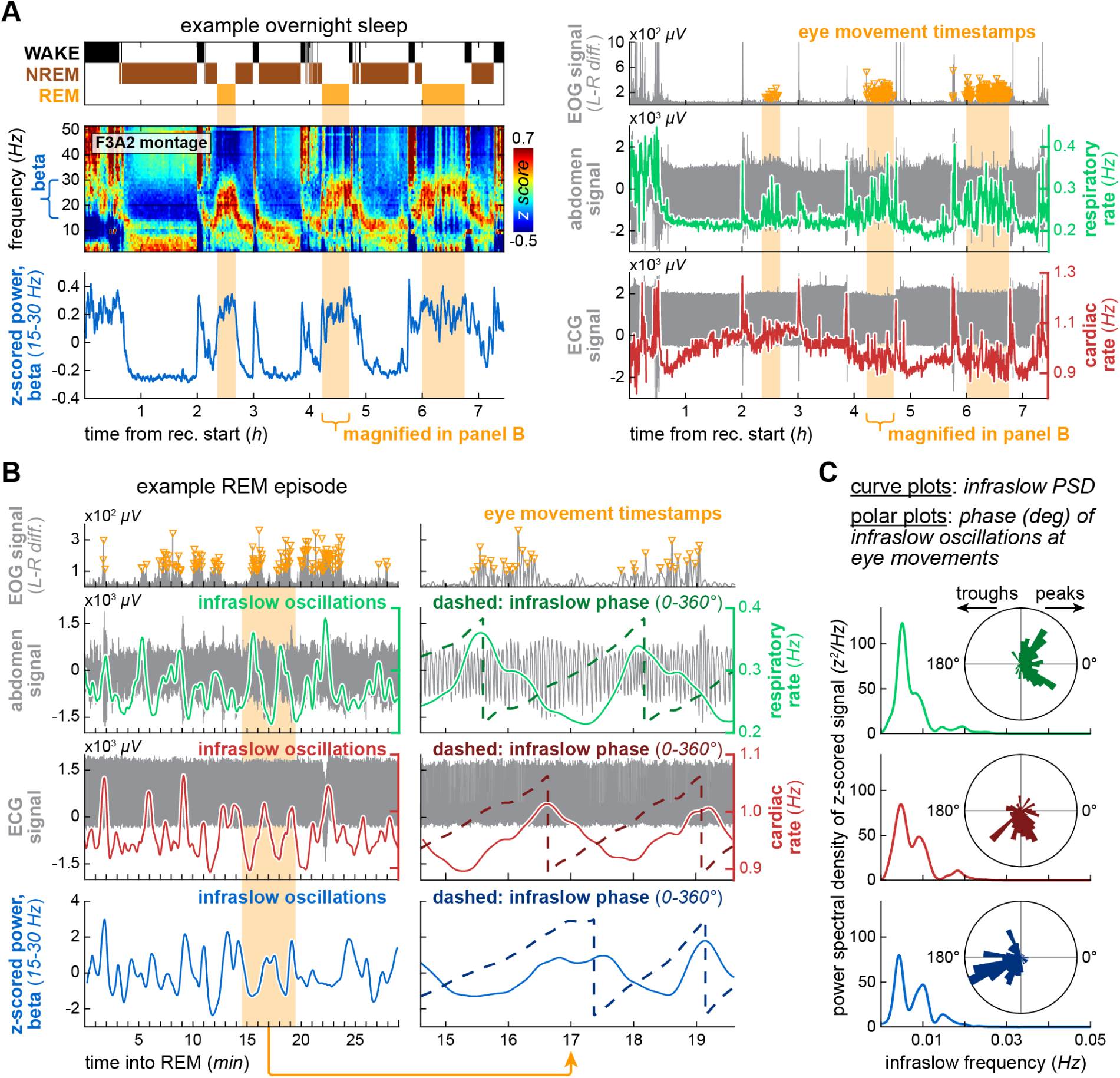
REM epochs and infraslow physiology metrics. (A) Representative overnight sleep. Left: REM epochs in the hypnogram are aligned with increased EEG beta activity, shown as a spectrogram and corresponding z-scored beta power curve. Right: same REM epochs, aligned with oculomotor activity and periods of increased respiratory and cardiac rate variability. Raw PSG signals are shown in gray, and derived data (detected eye movements, respiratory/cardiac rates) are shown in different colors. (B) Magnified view of the second REM episode in panel A. Left: full REM episode timeline, emphasizing oculomotor activity and physiological fluctuations on the timescale of minutes, i.e., infraslow rhythms. Right: further magnification illustrating the temporal alignment between oculomotor activity (timestamps) and infraslow rhythms (solid curves). Temporal shifts (phase offsets) among respiratory rate, cardiac rate, and beta power oscillations are illustrated by their Hilbert-transformed traces (dashed). (C) Infraslow periodicity and phase relationships from the same example REM episode are quantified by power spectral density and polar plots, respectively. All three physiological oscillations showed infraslow power peaks (∼0.005-0.01 Hz in this example) but differed in their phase alignment to eye movements. These patterns were generally consistent across participants, replicated our previous work [29], and were examined here for associations with neuropsychiatric and sleep health variables (Figures 3-5).

Using these methods, REM epochs scored by PSG technicians were plotted alongside beta activity, EOG activity, and periods of increased variability in cardiorespiratory rates (**Figure 2A**). This allowed us to verify and, when necessary, correct the sleep scoring by identifying isolated WAKE labels within otherwise continuous REM episodes. Discordant WAKE labels were then manually reclassified as REM, minimizing artifactual REM episode fragmentation. This data cleaning step was important for the infraslow periodicity analysis described next.

### REM infraslow physiology metrics

After REM epoch verification, individual REM episodes were processed as illustrated in **Figure 2B**. Like the methods above, the *findpeaks* function was used to detect eye movements, respiratory cycles and heartbeats. In particular, respiratory cycles and heartbeats were processed as follows: (1) their timestamps were counted in 1-s bins using *histcounts*; (2) each histogram was upsampled (interpolated) to the original sampling rate of 200 Hz, thus matching the length and temporal resolution of the corresponding REM episode; and (3) each upsampled histogram was smoothed (30-s moving mean followed by 30-s Gaussian smoothing), transforming them into respiratory and cardiac rate oscillations (**Figure 2B**). In turn, EEG beta power oscillations were derived from wavelet spectrograms (15-30 Hz frequency range, 1-Hz frequency resolution, 40 cycles) at the original sampling rate of 200 Hz, unlike the 1-s-binned spectrogram shown in **Figure 2A**. Each wavelet spectrogram was averaged across frequency bins, and infrequent outlier values in the resulting beta power trace (attributable to movement noise from REM sleep twitches) were filled by linear interpolation using percentile-based thresholds (*filloutliers* function, 0-95 percentile range) prior to smoothing (30-s moving mean followed by 30-s Gaussian smoothing). This yielded beta power oscillations like those obtained for respiratory and cardiac rates (**Figure 2B**).

All three oscillation types (respiratory rate, cardiac rate, beta power) were detrended per REM episode by subtracting their own 5-min moving averages and then z-scored, resulting in zero-centered semi-regular sinusoids amenable to spectral and phase analysis within the infraslow frequency band (**Figure 2B-C**). Before spectral analysis, REM episode signals were concatenated within each participant, so that summary spectral metrics represented individual participants rather than individual REM episodes. Then, power spectral density curves were generated using the *pwelch* function (10-min window, 50% overlap, 0-0.05 Hz frequency range, frequency resolution of 5×10^-5^ Hz) for each of the three infraslow oscillation types, per participant (**Figure 2B-C**). Analysis of individual, non-concatenated REM episodes was outside our scope but is a direction for future studies (see Discussion). For phase analysis, each infraslow oscillation was Hilbert-transformed into phase values (radians). Phase values indexed by eye movement timestamps were then extracted for polar plotting and circular statistics (**Figure 2B-C**). According to our previous work in healthy participants [29], oscillations in respiratory rate, cardiac rate and EEG beta power are quantifiable within the infraslow frequency band, but occur at different phases relative to eye movements. Here, these results were replicated in another sample of participants (**Figure 2C**) and further analyzed for correlations with neuropsychiatric and sleep health variables (**Figures 3-5**).

**Figure 3.**
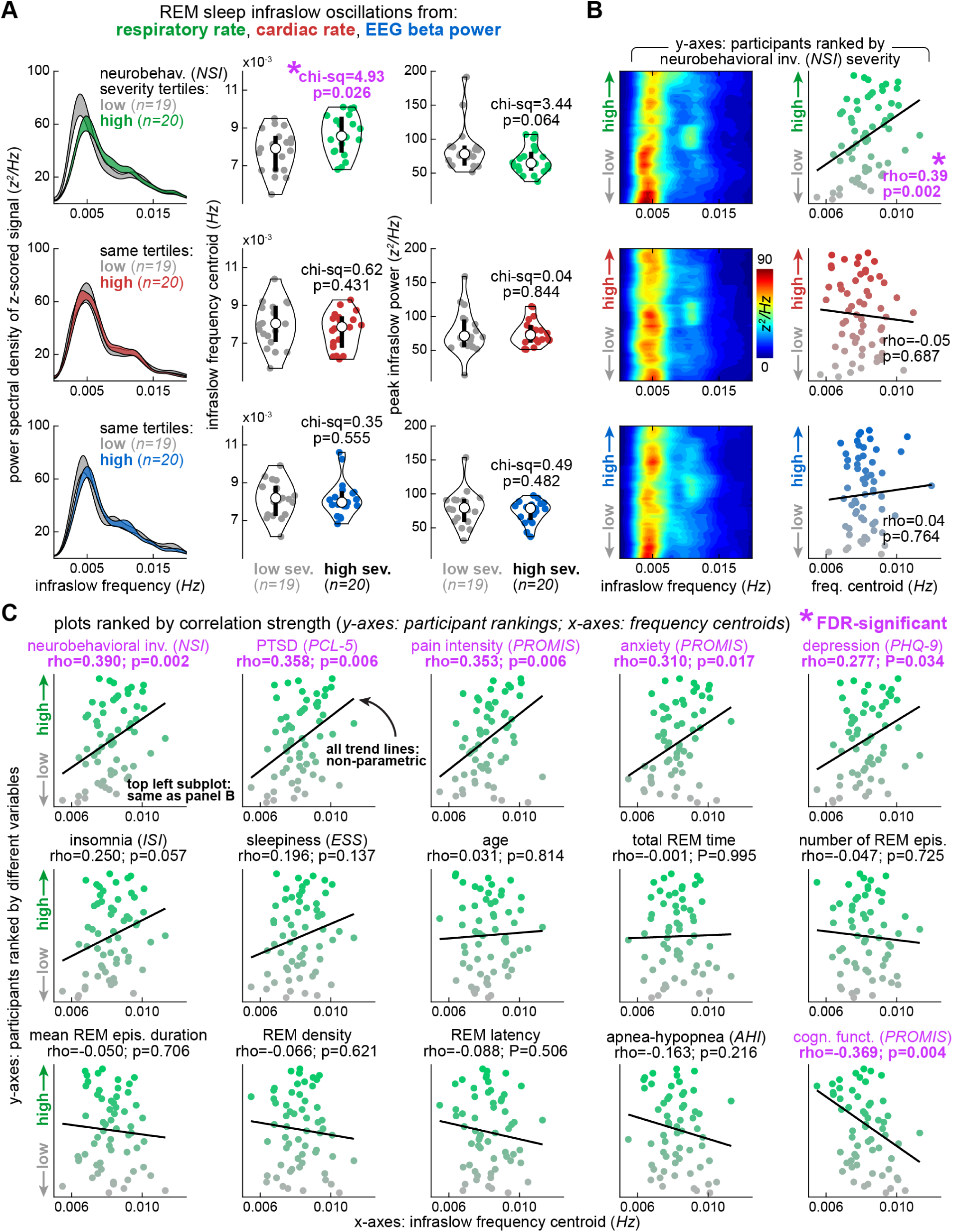
REM respiratory rate oscillations showed higher infraslow frequency with neuropsychiatric severity. (A) Left: infraslow power spectral density curves comparing low- and high-severity groups for a representative symptom domain: neurobehavioral symptom inventory (NSI). X-axes: infraslow frequency; y-axes: infraslow power. Shaded areas depict ± standard error. Low-severity curves are gray, and high-severity curves are colored differently per physiological domain. Violin distributions: infraslow frequency centroid (center column of subplots) and peak infraslow power (right column). Each datapoint represents one participant, unlike the standard error curves on the left. REM respiratory rate infraslow frequency was significantly higher in the high-severity group (top center), consistent with the qualitative group difference in the spectral density curves (top left). (B) All 59 participants are shown, ranked by NSI severity on the y-axes. Heatmaps show spectral density as a color scale, and scatter plots show corresponding Spearman correlations between infraslow frequency centroids and participant rankings. Higher REM respiratory rate infraslow frequency was associated with higher NSI severity, as shown qualitatively in the heatmap and quantitatively in the correlation. (C) Expanded correlation grid with participants ranked by different neuropsychiatric and sleep quality variables on the y-axes. REM respiratory rate infraslow frequency is shown on all x-axes. See supplements for additional scatter plots (**Figure S1-S4**). Subplots are ordered by Spearman rho, from strongest positive at the top left (NSI ranking on y-axis, same as panel B) to strongest negative at the bottom right. After false discovery rate (FDR) correction of the p-values in this panel, significant correlations were found between infraslow frequency and a subset of neuropsychiatric variables. Trend lines illustrate non-parametric fits, whose direction is negligible when rho is near zero.

Complementarily, eye movement timestamps were converted into continuous time series using the same binning, interpolation, smoothing, detrending and z-scoring procedures described above for respiratory and cardiac rates. The resulting curves (here referred to as eye movement rate oscillations) were then analyzed using the same infraslow spectral and phase methods (**Figure 4**, **Figures S1-S2**). Since eye movement timestamps were used to both generate and phase-index the eye movement rate oscillations, strong in-phase alignment was expected for this analysis, which served as additional control for comparison with other infraslow oscillations (**Figure 4**, **Figures S1-S2**). This complementary analysis also placed eye movements, i.e., one of the bases for phasic/tonic REM classification, within the proposed infraslow rhythm framework, allowing to test whether symptom associations extended to REM temporal structure as a whole or its specific physiological aspects.

**Figure 4.**
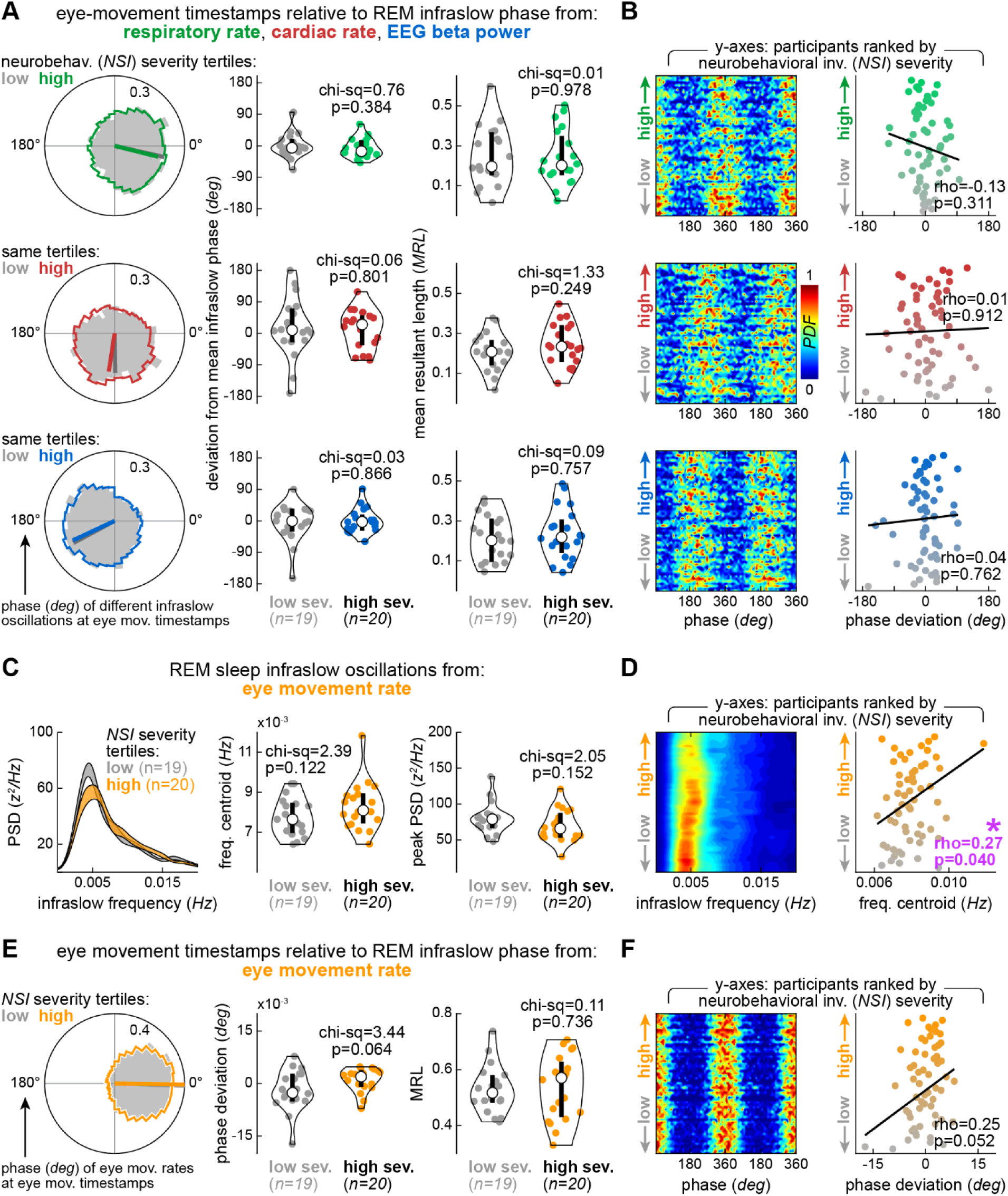
Eye movement timing within REM infraslow physiology showed limited symptom associations. (A) Polar plots: timing of rapid eye movements relative to the three types of REM infraslow cycles. Eye movements occurred preferentially during periods of high respiratory rate (∼0°), low EEG beta power (∼180°), and intermediate cardiac rate (∼270°), indicating phase-shifted infraslow physiology across brain and body and replicating our previous work [29]. These phase relationships were similar between participants with low and high neurobehavioral symptom inventory (NSI) severity, differentiated by colors in these polar plots. NSI is shown here as a representative symptom domain, as in Figure 3. Violin distributions: comparisons between the same NSI severity groups in terms of phase deviation (center column of subplots; see Methods) and mean resultant length, or MRL [52] (right column). Each datapoint represents one participant, unlike the aggregated circular distributions on the polar plots. No significant group differences were observed for any physiological domain or phase metric. (B) All 59 participants are ranked by NSI severity on the y-axes. Heatmaps show phase distribution as a color scale, and scatter plots show corresponding Spearman correlations between phase deviations and participant rankings. No significant correlations with NSI severity were observed. (C) Eye movement timestamps were converted into eye movement rates and analyzed as infraslow oscillations using the same data processing applied to respiratory and cardiac rates (see Methods). Infraslow power spectral density curves were then generated comparing low and high NSI severity. Shaded areas depict ± standard error. Violin distributions: comparisons between the same NSI severity groups in terms of infraslow frequency centroids (center column of subplots) and peak infraslow power (right column) from eye movement rate oscillations. (D) Heatmaps show spectral density as a color scale, and scatter plots show corresponding Spearman correlations between infraslow frequency centroids (x-axes) and participant rankings (y-axes). The infraslow frequency of eye movement rate oscillations increased with higher NSI severity, similar to the results from REM respiratory rate, but see Discussion. (E-F) Phase analyses like panels A-B, now using eye movement rate as the underlying infraslow rhythm. No significant NSI group differences or correlations were observed, but see supplements (**Figure S1-S2**) and Discussion. Trend lines illustrate non-parametric fits, whose direction is negligible when rho is near zero.

### Statistical analysis and correlations

The distributions for age and health variables (**Figure 1B**) are statistically summarized in **Table 1**, which includes descriptive statistics (median and interquartile range) and normality test results (Anderson-Darling). Most variables (10 of 15) showed evidence against normality (**Table 1**). To account for this distributional heterogeneity, nonparametric statistical methods were used throughout the study, as described next.

For infraslow rhythm metrics, spectral density curves like those in **Figure 2C** were summarized as infraslow frequency centroid (in Hz) and infraslow peak power (in z^2^/Hz). In turn, circular distributions like those in **Figure 2C** were summarized as mean resultant length (MRL [52]) and deviation from mean phase (in degrees). The latter was calculated as the signed circular difference between each participant’s mean phase and the mean phase across participants, wrapped to the -180° to 180° range. Thus, four infraslow rhythm metrics were obtained across three physiological domains (respiratory rate, cardiac rate, beta power), for a total of 12 metrics per participant.

These infraslow physiology metrics were initially compared between participant groups (**Figure 3A** and **Figure 4A**) and subsequently examined through correlation analyses (**Figure 3B-C**, **Figure 4B-C** and **Figure S1-S2**). For group comparisons, participants were divided into severity tertiles based on the neurobehavioral symptom inventory (NSI) scale, which was selected as a representative symptom domain for this initial analysis. The lowest and highest symptom severity groups were then statistically compared using Kruskal-Wallis test, and chi-square and p-values were reported along with their subplots (**Figure 3A** and **Figure 4A**). Then, as shown in the scatter plots of **Figure 3B-C** and **Figure 4B-C** (supplemented in **Figure S1-S2**), Spearman correlations were calculated between different infraslow physiology metrics (x-axes) and participant rankings (y-axes), with rankings derived from neuropsychiatric and REM architecture variables. Spearman rho and p-values were reported along with each scatter plot, including highlights indicating correlations that remained significant after false discovery rate correction (FDR; *mafdr* function). FDR correction was conducted separately for each set of p-values in each figure. Trend lines were also included in each scatter plot, representing non-parametric fits (Theil-Sen) shown only for visualization. A supplementary analysis (**Figure S3-S4**) was conducted replacing infraslow physiology metrics by more conventional metrics: basal respiratory and cardiac rates, calculated as median rates (Hz) per participant, across REM episodes. The rationale was to examine the added value of infraslow rhythms as symptom correlates relative to basal cardiorespiratory rates, as these basal rates require less signal processing and therefore would be preferable if equally informative.

For a full view of correlations, Spearman rho matrices were generated using all variables, including neuropsychiatric scores, conventional REM metrics, REM infraslow physiology metrics, and basal REM cardiorespiratory rates (**Figure 5A**). To group variables according to correlation strength, hierarchical clustering was performed using correlation-based distance, defined as 1 minus the absolute Spearman rho value. Average linkage was used to generate a hierarchical tree, and a cluster cutoff was determined based on the largest increase in linkage distance (dashed line; **Figure 5A**). At this cutoff, seven intercorrelated variables were initially found to form a cluster. Then, for visualization, the correlation matrix was reordered according to dendrogram leaf order; within the constraints of the tree, branches were oriented to place the detected cluster and its strongest correlates toward the top of the matrix (**Figure 5A**). Correlation coefficients are displayed as a rho matrix (see color scale), and matrix-wide FDR-corrected statistical significance is indicated by asterisks (**Figure 5A**).

**Figure 5.**
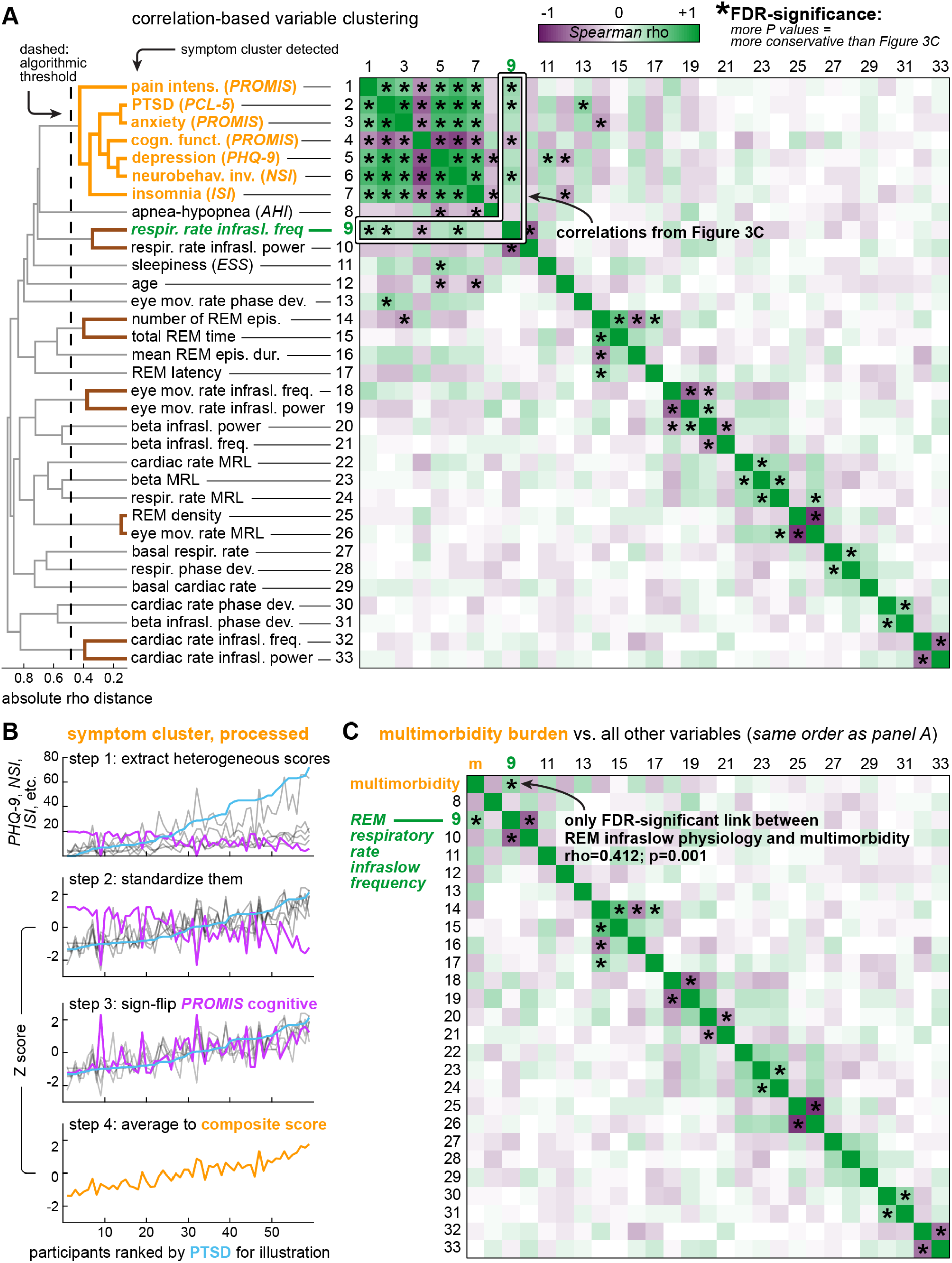
Correlation matrix revealed a symptom cluster associated with a specific REM infraslow rhythm. (A) Spearman correlation matrix across all variables, including neuropsychiatric, sleep quality and REM physiology metrics, for a total of 33 metrics per participant. The matrix is color-coded by Spearman rho, with asterisks indicating correlations that remained significant after false discovery rate (FDR) correction. Variable pairs corresponding to the scatter plots in Figure 3C are highlighted (see white outlines), contextualizing such variable pairs within the full correlation structure. As shown by the dendrogram on the left, variables were ordered by hierarchical clustering based on absolute Spearman rho distance (see Methods). The dashed line indicates an objective cutoff (largest increase in linkage distance) which in turn defined a cluster of seven intercorrelated symptoms, interpreted here as a multimorbidity burden (see variables highlighted in orange at the top). (B) This cluster of seven intercorrelated variables was further processed into a composite score per participant by standardizing the heterogeneous symptom scales into z-scores, sign-flipping PROMIS cognitive function (the only scale for which lower values indicated greater severity), and averaging the z-scores. This yielded one multimorbidity burden value per participant. (C) Reduced correlation matrix generated by replacing the individual variables in the symptom cluster with the composite multimorbidity score. All other variables are ordered as in panel A. REM respiratory rate infraslow frequency was the only REM infraslow periodicity metric showing an FDR-significant correlation with the composite symptom score.

Finally, the same methods were used to generate a reduced correlation matrix, in which the symptom cluster variables (**Figure 5A**) were summarized into a composite score (**Figure 5B-C**) representing multimorbidity burden. To calculate this composite score, the different scales within the symptom cluster were z-scored (**Figure 5B**). Cognitive function (PROMIS), in particular, was sign-flipped after z-scoring due to its opposite direction relative to the other scales (**Figure 5B**). Z-scored variables were then averaged into the composite score (**Figure 5B**), followed by repetition of the correlation methods described above (**Figure 5C**).

## Results

As described in the Methods section, we curated a retrospective PSG and symptom survey dataset to determine whether infraslow physiology during REM sleep is associated with neuropsychiatric symptom severity. The cohort consisted of 59 male participants with normal-to-mild apnea-hypopnea, sufficient REM sleep for infraslow physiology analyses, and broad distributions of age, sleep architecture variables, and neuropsychiatric symptom scores (**Figure 1**). We then examined REM infraslow rhythms across three physiological domains: respiratory rate, cardiac rate, and EEG beta power (**Figure 2A**). These were finally transformed into zero-centered oscillations for two quantitative approaches (**Figure 2B-C**): infraslow spectral analysis and infraslow phase analysis – this latter was performed relative to rapid eye movements. We next report results for infraslow spectral and infraslow phase analyses (**Figures 3-4**).

### REM respiratory rate oscillations showed higher infraslow frequency with neuropsychiatric severity

**Figure 3A** depicts spectral analysis for REM respiratory rate, cardiac rate, and beta power oscillations, with one oscillation type per subplot row. The left column of subplots shows power spectral density curves comparing two groups of participants: the lowest and highest tertiles of neurobehavioral symptom inventory (NSI) severity, with shaded areas representing ± standard error. In this initial qualitative comparison, group separation was apparent only for REM respiratory rate infraslow oscillations, with the spectral curve slightly shifted toward higher infraslow frequencies in the high severity group. This effect was statistically confirmed by the infraslow frequency centroid analysis in the second column of subplots (**Figure 3A**), where participants are represented by data points in the violin distributions. According to the data, REM respiratory rate oscillations showed higher frequency in the high-compared to low-severity group. Infraslow power was marginally (non-significantly) lower in the high-severity group (**Figure 3A**, right column of subplots), suggesting that infraslow frequency was more sensitive to NSI severity than infraslow power, at least for REM respiratory rate oscillations.

**Figure 3B** depicts data from all participants, sorted using a single NSI severity ranking, fixed across y-axes. The heatmaps in **Figure 3B** show spectral density data like those in the curve graphs of **Figure 3A**, with warmer colors indicating frequency bins with stronger power. A subtle tilt toward higher infraslow frequencies for REM respiratory rate oscillations can be noticed as NSI severity increases (**Figure 3B**). The top scatter plot in **Figure 3B** quantifies this trend through a significant Spearman correlation, showing that the higher the NSI severity, the higher the infraslow frequency centroid for REM respiratory rate. No significant correlations were observed for the infraslow frequency of REM cardiac rate or beta power oscillations. Together with **Figure 3A**, these results suggest an association between NSI severity and the infraslow frequency of REM respiratory rate oscillations.

**Figure 3C** shows the same infraslow frequency quantification for REM respiratory rate across all measured neuropsychiatric and basic REM variables (see subplot titles). The subplots themselves were sorted by Spearman rho values, from strongest positive at the top left to strongest negative at the bottom right. As indicated by FDR-significant p-values, infraslow frequency centroids for REM respiratory rate oscillations were correlated with neurobehavioral symptoms, cognitive difficulties, and scores for PTSD, pain, anxiety and depression, mentioned here in descending order of correlation magnitude, irrespective of direction.

Thus, **Figure 3** as a whole progresses from a group-level analysis of a representative symptom domain (**Figure 3A**) to a broader correlation grid across participant rankings (**Figure 3C**). As a supplementary analysis, **Figure S1** compares these respiratory rate infraslow frequency correlations with analogous correlations for cardiac rate and EEG beta power, neither of which yielded FDR-significant results. Furthermore, no correlations were observed when analyzing basal REM respiratory or cardiac rates (**Figures S3-S4**), suggesting that these basal rates, measured directly in Hz, were not sensitive to the neuropsychiatric variables studied in this dataset. Therefore, among REM physiology metrics, the infraslow frequency of respiratory rate oscillations showed the broadest associations with neuropsychiatric symptoms.

### Eye movement timing within REM infraslow physiology showed limited symptom associations

**Figure 4A-B** is organized similarly to **Figure 3A-B**, again using NSI severity for initial context. The polar plots in **Figure 4A** depict phase relationships between rapid eye movements and REM infraslow oscillations from the three physiological domains, aggregated across all REM episodes from all participants (see **Figure 2B-C** for a representative REM episode). Rapid eye movements occurred preferentially around the peaks of respiratory rate cycles (phase concentration around 0°), troughs of beta power cycles (around 180°), and an intermediate phase of cardiac rate cycles (around 270°), replicating our previous work [29]. The violin distributions (**Figure 4A**) quantify phase deviations for individual participants relative to the mean phase patterns shown in the polar plots, as well as MRL, a measure of phase concentration irrespective of phase angle [52]. No significant differences were observed between participant groups for either measure (**Figure 4A**), suggesting that phase relationships between eye movements and underlying infraslow oscillations were not sensitive to NSI severity. The heatmaps and scatter plots in **Figure 4B** likewise showed no associations between the phase deviation metric described above and NSI severity, regardless of physiological domain, whether respiratory rate, cardiac rate or EEG beta power. Expanded correlation analyses between phase deviation and all participant rankings also yielded no associations across these three physiological domains (**Figure S2**).

To examine the oculomotor component of REM infraslow physiology more directly, eye movement timestamps were processed into continuous time series using the same procedures applied to respiratory and cardiac rates (i.e., binning, interpolation, smoothing, detrending and z-scoring; see Methods). This yielded eye movement rate oscillations amenable to infraslow spectral analysis, as shown in **Figure 4C**. The high NSI severity group showed a shift toward higher infraslow frequencies and reduced peak prominence in the spectral curve, qualitatively resembling the patterns observed in respiratory rate (**Figure 3A**). Corresponding trends in infraslow frequency and power did not reach significance (**Figure 4C**). When analyzed as a function of participant rankings, eye movement rate infraslow frequency did correlate significantly with NSI severity (**Figure 4D**), but this association was weaker than the corresponding one for respiratory rate (**Figure 3B**). In addition to NSI severity, pain intensity was the only other correlate of eye movement rate infraslow frequency (**Figure S1**): a smaller range of neuropsychiatric associations than that observed for REM respiratory rate.

Finally, eye movement rate oscillations were analyzed for phase relationships with the same eye movement timestamps from which they were derived, for additional comparison with the other three types of infraslow oscillations (respiratory, cardiac, and EEG beta activity). As expected, eye movements showed strong phase preference to the peaks of their own rate oscillations (**Figure 4E**). Phase deviations occupied a much narrower range (-15° to 15°, approximately) than other similar analyses, as shown by both the violin plot and scatter plot of **Figure 4E-F**. Nonsignificant trends toward positive correlations between phase deviation and NSI severity were observed (**Figure 4E-F**), but these trends were not interpreted further, given the narrow phase deviation range and the built-in dependence between eye movement rates and eye movement timestamps. MRL also showed no significant difference between NSI severity groups (**Figure 4E**). In a supplementary analysis (**Figure S2**), phase deviation for eye movement rate infraslow oscillations correlated only with PTSD and anxiety scores, thus showing fewer symptom associations than respiratory rate infraslow frequency (**Figure S1**).

Therefore, according to **Figures 3-4**, neuropsychiatric symptom associations were not uniformly expressed across REM infraslow physiology. For example, if symptom severity were reflected in a synchronized manner across REM infraslow rhythms, then respiratory and eye movement rates might be expected to show comparable neuropsychiatric associations, given their in-phase temporal relationship (see polar plots of **Figure 4A,E**). However, these associations only partially overlapped, with respiratory rate oscillations showing greater symptom sensitivity. Next, these findings are situated within the full correlation structure across all analyzed variables.

### Correlation matrix revealed a symptom cluster associated with a specific REM infraslow rhythm

**Figure 5A** shows a comprehensive panel of all variables: their pairwise correlations (Spearman rho matrix on the right), and their correlation structure as revealed by hierarchical clustering (dendrogram on the left). As explained in the Methods section, hierarchical clustering reordered the correlation matrix based on the absolute Spearman rho distance among variables, allowing strongly correlated variables to cluster together, regardless of direction. Thus, a data-driven cutoff in the hierarchical tree, reflecting the largest increase in linkage distance, revealed a cluster of intercorrelated symptoms (**Figure 5A**, see dashed line and highlighted variables).

The symptom cluster appears as a block of FDR-significant correlations at the top left of the matrix (**Figure 5A**), particularly among variables indexed 1-7 after dendrogram-based reordering, namely: pain intensity, PTSD, anxiety, cognitive function, depression, neurobehavioral symptoms, and insomnia. This puts the findings shown earlier in **Figure 3C** into perspective: REM respiratory rate infraslow frequency indeed showed correlations with individual symptom scales (**Figure 5A**, see highlighted variable pairs on the correlation matrix), but these associations seemed embedded within a broader multimorbidity burden, as elaborated later (**Figure 5B-C**).

Other than the symptom cluster, the infraslow frequency of REM respiratory rate oscillations was negatively correlated with their own infraslow power (**Figure 5A**). Despite this frequency-power anticorrelation, REM respiratory rate infraslow power was not associated with any variable in the symptom cluster after FDR correction. Similar frequency-power anticorrelations were observed for cardiac rate, eye movement rate and beta power, but none showed symptom associations either (**Figure 5A**). Beyond these main patterns, isolated significant correlations were observed elsewhere in the matrix, including a positive correlation between depression and sleepiness, and a negative correlation between anxiety and number of REM episodes (**Figure 5A**). Because these associations were sparse and did not form a cohesive pattern involving neuropsychiatric symptoms and REM infraslow physiology, they were considered outside the scope of this study and were not interpreted further.

As suggested above, the intercorrelations within the symptom cluster seemed to indicate a multimorbidity burden, prompting us to test whether REM respiratory rate infraslow frequency covaried more strongly with such burden than with specific domains/disorders. To investigate this, the different numerical scales within the symptom cluster were standardized as z-scores (**Figure 5B**). When plotted across ranked participants, these z-scores revealed similar upward trajectories, except for PROMIS cognitive function, which followed the opposite direction and was therefore inverted (sign-flipped) (**Figure 5B**). Once aligned in the same direction, the z-scores were averaged into a single composite score by calculating the mean across all seven variables in the symptom cluster (**Figure 5B**). A reduced correlation matrix was finally generated using the composite score instead of the individual variables in the symptom cluster. Consistent with the full correlation matrix in **Figure 5A**, REM respiratory rate infraslow frequency was the only infraslow rhythm metric showing FDR-significant correlation with the composite symptom score (**Figure 5C**). This correlation was positive, indicating that the higher the multimorbidity burden, the higher the infraslow frequency of REM respiratory rate oscillations (**Figure 5C**).

The correlation between REM respiratory rate infraslow frequency and the composite symptom score is further illustrated in **Figure 6A**. Spearman rho and p-values were included in **Figure 6A** (rho = 0.412, p = 0.001) for comparison with the statistics reported earlier in **Figure 3C** (rho ≤ 0.390, p ≥ 0.002). The conclusion from these rho value comparisons is that the composite multimorbidity score showed a stronger association with REM respiratory rate infraslow frequency (**Figure 6A**), exceeding correlations with individual symptom domains such as NSI, cognitive function, PTSD, pain, anxiety and depression (**Figure 3C**). These findings expand our previously proposed model of REM microstructure [29] (**Figure 6C-D**), and raise questions for future studies on REM physiology and mental health.

**Figure 6.**
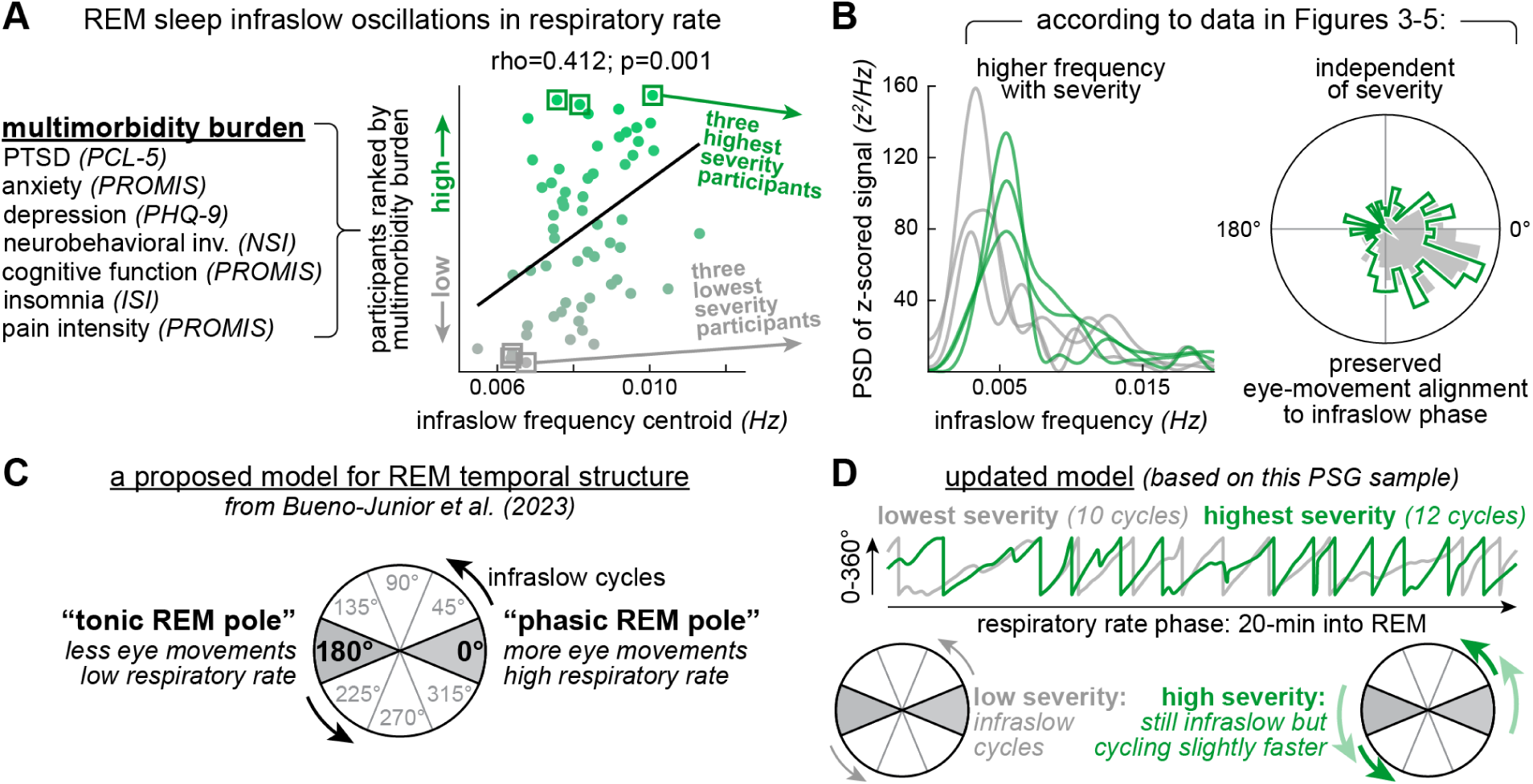
Main findings and proposed model for REM temporal structure. (A) Left: multimorbidity symptoms defined in Figure 5. Right: correlation between multimorbidity severity (y-axis) and infraslow frequency from REM respiratory rate oscillations (x-axis). This correlation strength (rho=0.412; p=0.001) exceeded all those reported in Figure 3. Trend line: non-parametric, for illustration. (B) The three lowest- and three highest-ranked participants in the scatter plot in panel A are further illustrated by their respiratory rate power spectra (curve graph) and circular distributions relative to eye movements (polar plot). This qualitative view summarizes the positive correlation between symptom cluster severity and REM respiratory rate infraslow frequency, while no change was detected in the phase alignment of respiratory rate cycles with rapid eye movements. (C) Proposed model for REM temporal structure, first introduced in our previous work [29]. Respiratory and eye movement rates were initially suggested to cycle together at an infraslow timescale between phasic-like and tonic-like poles. (D) Updated model based on the present findings. Time series: REM respiratory-rate infraslow phase from the lowest- and highest-severity participants highlighted in panels A-B. These real-data epochs (20-min each) were obtained independently from each participant and are overlaid for illustration. The higher-severity participant shows two more infraslow cycles within this example 20-min period, consistent with the infraslow frequency differences in panels A-B. Polar plots: the proposed model for REM temporal structure is shown again, now incorporating the neuropsychiatric dimension. The present findings suggest physiologically selective symptom associations, with REM respiratory rate infraslow frequency providing a stronger readout of symptom severity than oculomotor activity. More research is needed to better characterize the coordination among these and other REM infraslow rhythms in relation to neuropsychiatric state.

## Discussion

This study examined whether REM infraslow physiology relates to neuropsychiatric severity in male veterans. Higher multimorbidity burden (pain intensity, PTSD, anxiety, cognitive function difficulties, depression, neurobehavioral symptoms, and insomnia) was associated with higher frequency in REM respiratory rate infraslow oscillations, identifying a potential new readout of neuropsychiatric symptoms in more diverse populations, as well as a physiological target for mechanistic studies in animal models of disease.

### Infraslow rhythmicity as a framework for REM physiology

The temporal structure within REM sleep is commonly described as alternating between two substates, phasic and tonic REM [3], distinguished most prominently in the literature by high versus low incidence of motor transients, respectively, but also by differences in sensory sensitivity and brain physiology [12–27]. This framework has guided research on REM-related physiological variations, emotion processing, memory consolidation, and neurodevelopment in both humans and animal models [23,53–61]. However, this binary phasic/tonic framework leaves two aspects of REM temporal structure underexplored: the periodicity with which phasic and tonic REM alternate, and the degree to which these substates are discretely separable. We propose a complementary view: phasic and tonic REM may represent poles within a minute-scale (i.e., infraslow) continuum spanning physiological signals across brain and body (**Figure 6C**). Specifically, our previous work analyzed REM in six mice and nine healthy humans [29]. In humans, we found that rapid eye movements occurred preferentially near respiratory rate peaks, EEG beta power troughs, and an intermediate phase of cardiac rate, all oscillating at infraslow frequencies. This suggested that oculomotor, autonomic, and brain dynamics during REM sleep are organized by a shared temporal structure, with different infraslow phases among brain/body signals [29]. The present study replicated these temporal relationships in a separate human sample (n=59) and additionally found that one particular aspect of REM, the infraslow frequency of respiratory rate oscillations, varies with neuropsychiatric multimorbidity (**Figure 6D**). Given the in-phase temporal relationship we observed between respiratory and eye movement rate oscillations, the latter were also examined for infraslow periodicity in the present study. The hypothesis was that both physiological aspects (respiratory and oculomotor) would be jointly associated with symptom severity, thus implying a broader alteration of REM temporal structure. However, REM respiratory rate oscillations, and more specifically their infraslow frequencies, emerged as the clearest correlate of neuropsychiatric state. This suggests that either: (1) neuropsychiatric state is not uniformly reflected across REM infraslow physiology; (2) respiratory rate provides a more sensitive symptom readout simply because abdominal plethysmography yields a cleaner signal compared to eye movement rates; or (3) a combination of both. Distinguishing among these possibilities merits further investigation.

Despite these intricacies, our findings add to growing evidence that sleep physiology exhibits infraslow rhythmicity. A comparative biology study (Bergel et al., 2026) [62] reported that sleep infraslow rhythms are evolutionarily conserved across reptiles and mammals. This phylogenetic framing is a recent development in a research line that began in the 2004-2016 period, when infraslow activity was initially quantified during human sleep, particularly NREM [7,63–65]. These early studies showed that K-complexes (NREM-typical EEG events) and neocortical-subcortical signal propagation (measured via functional imaging) are all organized at infraslow timescales during human NREM [7,63–65]. Subsequently, over the past decade, this literature on sleep infraslow rhythms has incorporated additional EEG patterns typical of NREM: sleep spindles and spindle-related sigma power in both humans [2,66–73] and rodents [2,74–84]. Specifically, human studies have examined infraslow oscillations in spindle incidence and sigma power in relation to sleep fragility, memory consolidation, and different neuropsychiatric conditions [2,66–71,73], whereas rodent studies have linked these NREM infraslow rhythms to locus coeruleus norepinephrine dynamics, hypothalamic/brainstem circuit activity, brain-body physiological fluctuations, neocortical interneuron activity and pontomedullary circuits regulating NREM-REM transitions [2,74–84]. Beyond spindle and sigma activity in the EEG, NREM infraslow rhythmicity has also been linked to coordinated monoaminergic and cholinergic modulation, as well as gastric rhythms and autonomic, vascular, cerebrospinal fluid, and glymphatic clearance dynamics, with implications for memory, sleep restoration, fatigue and neurodegenerative vulnerability [10,85–95]. Thus, NREM infraslow rhythms are increasingly viewed as a scaffold for brain-body rhythmicity.

REM sleep, in contrast, has only more recently been examined through this “infraslow lens”, so far represented by fragmented evidence from few studies. Neocortical interneuron activity in mice has been shown to oscillate at infraslow frequencies during NREM, as cited above, and according to the same work (Rolle et al., 2025) [84] this same infraslow rhythm persists into REM, albeit with lower amplitude. Elsewhere in the mouse brain, particularly in the midbrain and pons, neurons known to promote and suppress REM (REM-on/REM-off neurons) have additionally been shown to engage in opposing infraslow dynamics, potentially gating NREM-REM transitions (Lozano et al., 2026) [96]. And beyond state transitions, infraslow rhythms have been reported in rat hippocampal theta synchrony within REM itself, with neuronal firing across neocortical and thalamic areas phase-locked to these infraslow rhythms (Espinosa et al., 2026) [97]. Very similar findings had been reported in our own mouse experiments (Bueno-Junior et al., 2023) [29], including infraslow oscillations in hippocampal theta frequency during REM, as well as phase coupling of such oscillations to both cortical spiking and eye movements. These converging findings in rodents, along with the neuronal activity and state-transition reports cited above, begin to outline the underpinnings of REM infraslow microstructure. Thus far, these emerging studies suggest that infraslow rhythms across distributed brain circuits may contribute to, or at least reflect, the transition into REM and the organization of REM microstructure once the state is fully established.

Thus, more mechanistic research in laboratory animals is now needed to determine whether REM infraslow rhythms are causal drivers of REM microstructure, passive readouts, or both. Indeed, our work in humans captured phase-shifted infraslow rhythms from PSG body signals, which provides useful phenomenology but limited insights into neural substrates. A good starting point for mechanistic interrogation in rodents would include the same brain areas mentioned above (hypothalamus, brainstem, thalamus, hippocampus, neocortex) [2,78,80,93,96,97], and potentially the amygdala and related limbic circuits: a neural substrate jointly implicated in autonomic control, emotion regulation, and REM initiation/maintenance [98]. Furthermore, alterations in NREM infraslow-related measures have been linked to schizophrenia, autism spectrum disorders, post-infection chronic fatigue (long COVID), and amyloid accumulation (Alzheimer’s disease) [70–72,85,89,95], hence analogous alterations during REM may be testable in rodent models of disease, encouraged by our human data.

### Questions and future directions

Aside from these mechanistic speculations, this study raises a few caveats and also opportunities for developing sleep-based diagnosis. First, the symptom associations observed here were stronger for REM respiratory rate, weaker for eye movement rate, and absent in cardiac rate and EEG beta power. Why? Aside from signal quality differences among these readouts, could this mean that variations in their infraslow phase alignment patterns reflect distinct symptom profiles? Or could this heterogeneity among different infraslow rhythms merely reflect limited sample size? Second, with the participant cohort used here, we were able to extend our previous observations on REM infraslow physiology from healthy humans [29], but further interpretations are limited by the demographic composition of the present PSG sample, i.e., male veterans. Although this sample limits generalizability, it still provides a foundation for future REM microstructure studies using public datasets [99] and more representative civilian cohorts, including participants differing in gender, medication, and health conditions not studied here, such as REM sleep behavior disorder (RBD) and other forms of neurodegenerative risk [100–113]. Third, residual confounding by apnea-hypopnea cannot be ruled out, as hierarchical clustering placed AHI adjacent to the main group of intercorrelated symptoms, according to the data reported here. Still, AHI did not correlate significantly with the composite multimorbidity score or any respiratory rate metric, consistent with our data curation effort to minimize apnea in this PSG sample. Future studies could test whether these findings are replicated in cohorts recruited independently of respiratory complaints.

Fourth, this is a cross-sectional study, and therefore it did not capture longitudinal aspects of REM infraslow physiology metrics, like variations in these metrics over days or weeks. This is also not necessarily a limitation, as REM infraslow physiology remains a young field, with ample room for exploration across study designs. Thus, even within the constraints of a cross-sectional design, this study already shows that REM infraslow rhythms are worth investigating clinically, as they might carry symptom-relevant information. In addition, because the REM infraslow physiology metrics described here can be derived from cardiorespiratory signals, existing portable sleep monitoring devices could record these metrics, without fundamentally new hardware. This could enable longitudinal tracking of REM infraslow physiology metrics and associated dynamics (e.g., sleep deprivation, mood changes, treatment response) over weeks/months, provided that further PSG-based research confirms the clinical relevance of these metrics. Finally, future machine learning work could test whether REM infraslow physiology adds predictive value beyond conventional sleep quality metrics. Such models could be trained on variable sets with versus without REM infraslow physiology metrics, and then tested for their performance at classifying participants into symptom severity groups. Furthermore, individual REM bouts could be analyzed using these models, potentially revealing episode-to-episode REM dynamics that are obscured when data are summarized per participant, e.g., differences in infraslow periodicity between REM episodes occurring early and late in the night.

### Concluding remarks

The associations presented here between REM infraslow rhythms and neuropsychiatric multimorbidity add to emerging evidence that physiological variations within sleep states are relevant to health and disease. Further replication across larger and more diverse datasets is required to establish disease associations, but this and our previous work [29] already consolidate REM infraslow physiology as a new dimension of sleep quality. Given that REM infraslow rhythms are expressed widely across cardiorespiratory, oculomotor and brain activity, it is plausible that these rhythms may be relevant to sleep neurobiology, physiological phenotyping, and neuropsychiatric monitoring, which is a direction for future research.

## Acknowledgements

We thank the participants whose deidentified PSG and symptom survey data contributed to this study. We also thank our laboratory colleagues for helpful discussions. Generative artificial intelligence (U-M GPT) was used for polishing human-generated text and creating the icons in **Figure 1A**. All interpretations and conclusions expressed here are the responsibility of the authors and do not necessarily reflect the position or policy of the Department of Veterans Affairs, the National Institutes of Health, or the United States government.

## Data availability

The current version of the analysis code is available in the following GitHub repository: https://github.com/brendonw1/WatsonLabCode/tree/master/HumanREMinfraslow. Upon acceptance of this manuscript, the code will be archived in a DOI-issuing public repository. Deidentified, processed data will be made available upon reasonable request.

