## Supplementary material for "REM sleep infraslow rhythms correlate with neuropsychiatric symptom severity"

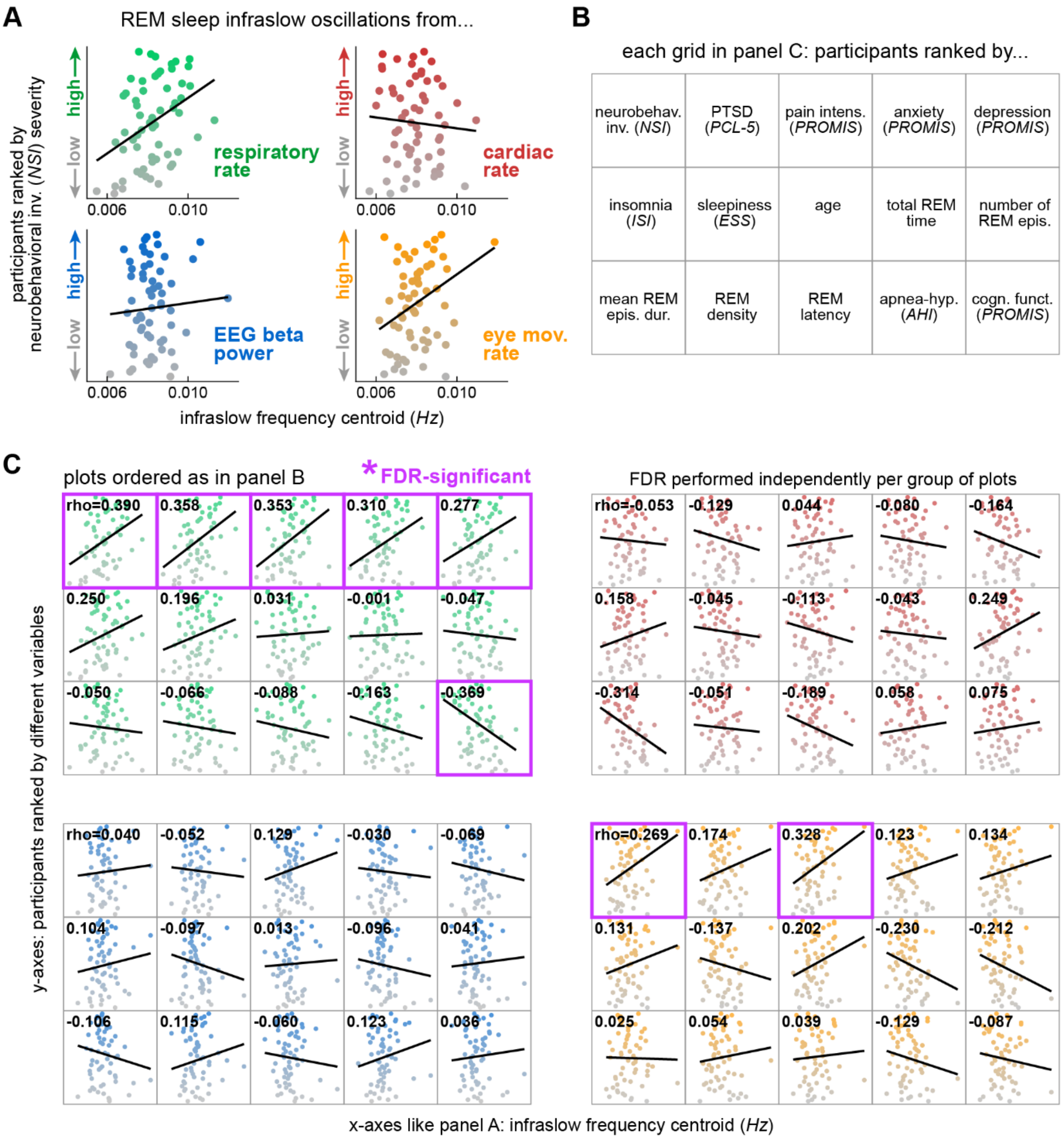


**Figure S1. REM sleep infraslow frequency associations across physiological and neuropsychiatric symptom domains**. (A) Scatter plots show Spearman correlations between infraslow frequency centroids from each physiological domain (infraslow oscillations in REM respiratory rate, cardiac rate, EEG beta power and eye movement rate) and participant rankings for neurobehavioral symptom inventory (NSI) severity, shown here as a representative symptom domain, as in **Figures 3-4**. Trend lines illustrate non-parametric fits (Theil-Sen). (B) Positional key identifying the y-axes at each subplot location in panel C (same subplot order as **Figure 3C**). (C) Correlation grids for each physiological domain. All x-axes show infraslow frequency centroids and the y-axes correspond to those in panel B. Spearman rho values are reported per subplot. False Discovery Rate (FDR) correction was applied separately for each physiological domain, and correlations that remained significant are highlighted.

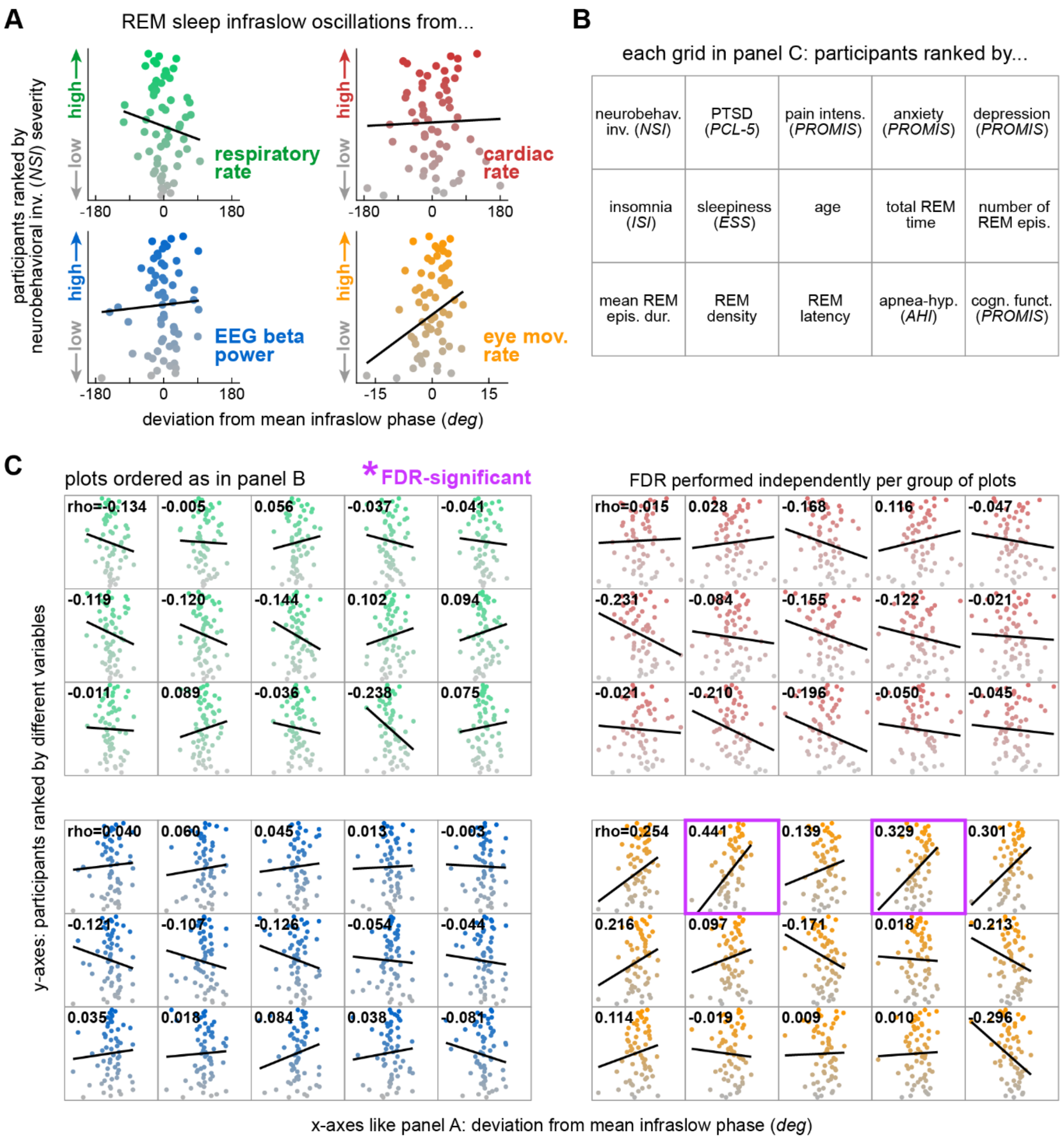


**Figure S2. REM sleep infraslow phase associations across physiological and neuropsychiatric symptom domains**. (A) Phase deviation was defined as the circular difference between each participant's mean eye movement-locked phase and the corresponding mean phase across participants. Scatter plots show Spearman correlations between this metric from each REM infraslow oscillation type (respiratory rate, cardiac rate, EEG beta power and eye movement rate) and participant rankings for neurobehavioral symptom inventory (NSI) severity, shown here as a representative symptom domain, as in **Figures 3-4**. Trend lines illustrate non-parametric fits (Theil-Sen). (B) Positional key identifying the y-axes at each subplot location in panel C (same subplot order as **Figure 3C**). (C) Correlation grids for each physiological domain. All x-axes show phase deviation and the y-axes correspond to those in panel B. Spearman rho values are reported per subplot. False Discovery Rate (FDR) correction was applied separately for each physiological domain, and correlations that remained significant are highlighted.

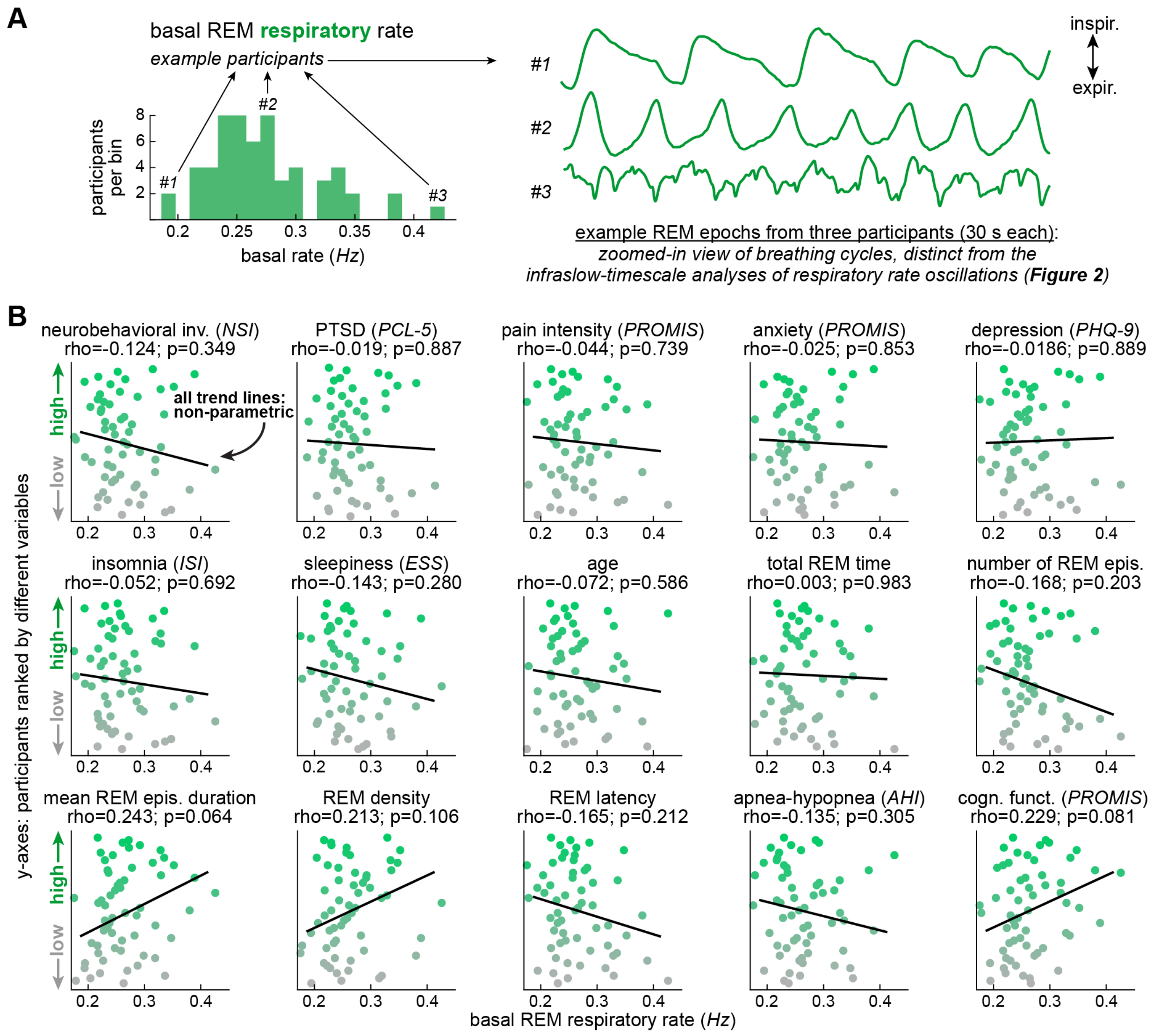


**Figure S3. No associations between basal REM respiratory rate and neuropsychiatric or sleep health variables**. (A) Distribution of participants across basal REM respiratory rate bins, like the distributions in **Figure 1B**. Arrows indicate bins in the lower, median, and upper regions of the distribution, from which three example participants were selected for the representative plethysmography traces on the right. The example traces (30 s each) illustrate individual breathing cycles in these three participants, supplementing the longer-timescale analyses in the main figures. (B) Scatter plots showing participants ranked by different neuropsychiatric and sleep quality variables on the y-axes. Basal REM respiratory rate is shown on the x-axes. Subplots are ordered as in **Figure 3C** for comparison. No significant correlations were observed for basal REM respiratory rate, in contrast to the associations involving REM respiratory rate infraslow frequency (**Figure 3C**). Trend lines illustrate nonparametric fits (Theil-Sen).

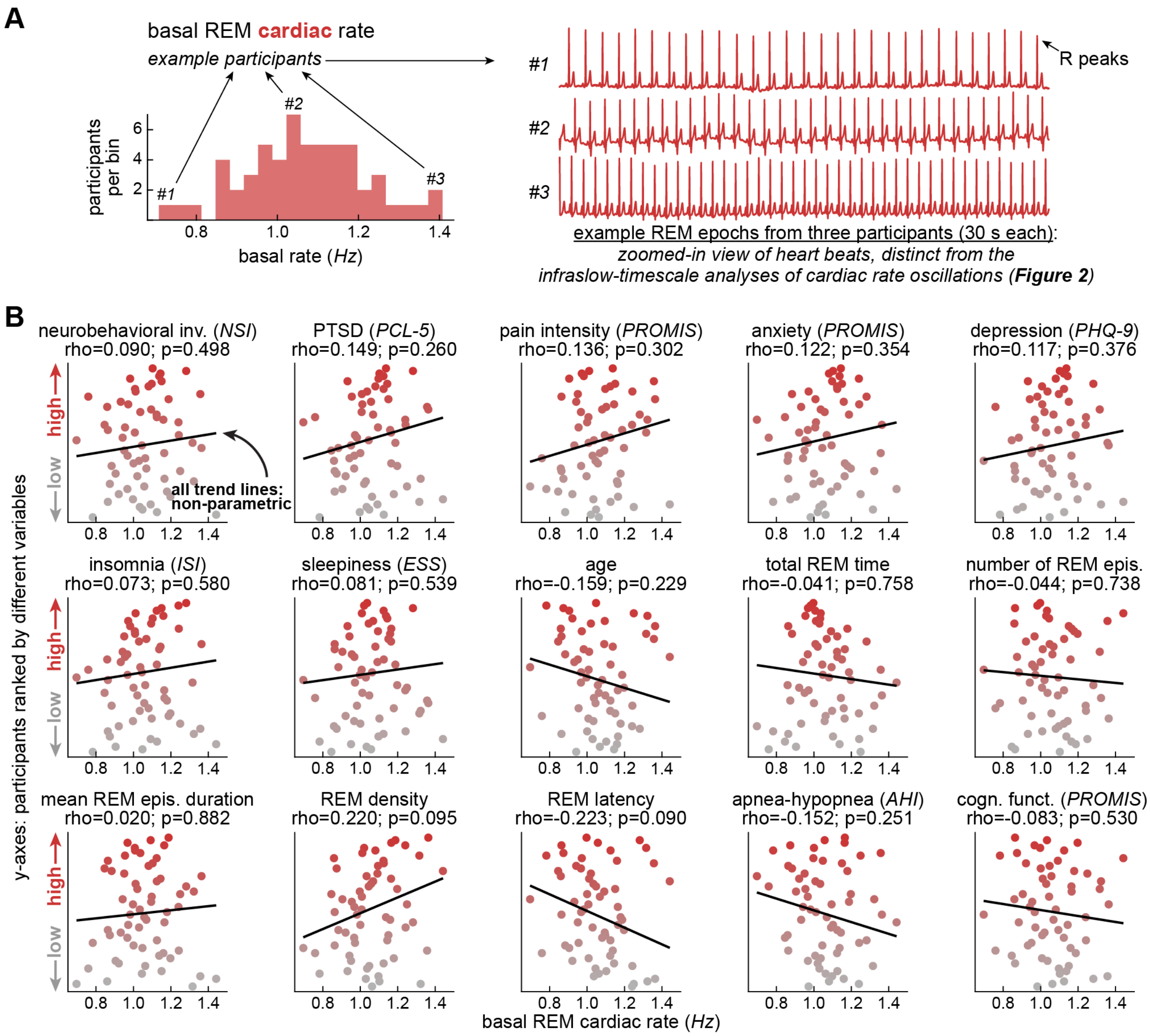


**Figure S4. No associations between basal REM cardiac rate and neuropsychiatric or sleep health variables**. (A) Distribution of participants across basal REM cardiac rate bins, like the distributions in **Figure 1B**. Arrows indicate bins in the lower, median, and upper regions of the distribution, from which three example participants were selected for the representative electrocardiographic traces on the right. The example traces (30 s each) illustrate individual cardiac cycles in these three participants, supplementing the longer-timescale analyses in the main figures. (B) Scatter plots showing participants ranked by different neuropsychiatric and sleep quality variables on the y-axes. No significant correlations were observed for basal REM cardiac rate, like **Figure S1** shown above. Trend lines illustrate nonparametric fits (Theil-Sen).
